# Mitochondrial Disease variation in healthy older adults: a genotype-phenotype assessment linking pathogenic variants and mitochondrial constraint

**DOI:** 10.64898/2026.06.24.26356498

**Authors:** Eloise Watson, Gordon Qian, Shyamsundar Ravishankar, Matthew Hobbs, Joseph Copty, Chenglong Yu, Sarah Kummerfeld, Christina Liang, Paul Lacaze, Ryan L. Davis, Carolyn M. Sue

## Abstract

Mitochondrial diseases (MDs) are caused by variants in the mitochondrial (mtDNA) or nuclear (nDNA) genome and encompass a diverse disease spectrum, whilst mitochondrial dysfunction more broadly is implicated in aging and neurodegeneration, with distinct and overlapping phenotypic features. Recent population genomic studies reveal pathogenic MD variation to be common in the population and somatic mtDNA variants accumulate from the seventh decade. Cumulative burden of mtDNA variation, quantified using mitochondrial genome constraint measures, may mediate mitochondrial dysfunction generally. However, the clinical relevance of incidentally identified variation for MDs, and of mitochondrial constraint measures for aging and neurodegeneration, is unclear.

We have quantified pathogenic mtDNA and nDNA variation, as well as measures of mitochondrial genome constraint in the Medical Genome Reference Bank (MGRB), a cohort of healthy older individuals. We evaluated association of identified pathogenic MD variants with clinical features relevant to MD across four domains including physical function, cognitive function, endocrine-metabolic function and mood. Associations of mitochondrial genome constraint with clinical measures of aging and neurodegeneration were also explored.

No significant associations between MD variants and phenotypes were identified, although surprisingly, mood measures appeared healthier for variant carriers compared to non-carriers. Summed mtDNA constraint showed significant inverse association with blood pressure and a trend toward inverse association with physical function. Measures of cognitive function did not demonstrate association with summed or mean mitochondrial genome constraint.

Pathogenic MD variants are relatively common in the population and may be carried through to old age in good health, emphasising the importance of clinical context for counselling. Mitochondrial genome constraint measures warrant further evaluation as a surrogate biomarker for mitochondrial genome quality and mitochondrial dysfunction.

## Introduction

Mitochondria are critical cellular organelles, responsible for generating the majority of cellular energy through oxidative phosphorylation (OXPHOS). Mitochondrial diseases (MDs) are a heterogeneous group of multisystem disorders resulting from pathogenic variants that impact mitochondrial function.(1, 2) Pathogenic variants occur in either the mitochondrial (mtDNA) or nuclear (nDNA) genomes, with nearly 400 disease-associated genes identified.(3–5) MDs can be challenging to diagnose in clinical practice, due to their complex genetic aetiology and broad spectrum of clinical features, ranging from mild, single-organ symptoms to life-threatening, multisystemic disease which may overlap with common disorders including diabetes and stroke.(6–9).

Due to the marked genetic and phenotypic heterogeneity of MDs, they are difficult to capture epidemiologically. Traditional epidemiological studies of MD prevalence have primarily drawn from clinical cohorts, with population risk estimates derived from pedigree analysis.(2, 10–22) Although these studies show variability in estimates, MD prevalence is commonly reported as 1 in 5,000 affected individuals,(12, 23) with around 1 in 4,000 individuals thought to carry a genetic variant placing them at risk.(23) However, early population-based studies examining the prevalence of one or a few mtDNA variants suggested a substantially higher carrier rate, (1 in 139 to 244 individuals) that has been replicated in multiple studies, possibly with clinical manifestations.(18–22) More recently, analysis of genetic reference cohorts has supported the conjecture that pathogenic MD variants are more common in the population than estimated from clinical cohorts.(24–27) However, prior studies suggest reduced variant penetrance in unselected populations.(28–31) Therefore, an understanding of variant penetrance and expression in clinically unselected carriers, compared to non-carriers and affected individuals, is critical to understand the clinical significance of this pathogenic MD variation.

Beyond monogenic MDs, mitochondrial dysfunction is also robustly linked to age-related and neurodegenerative processes through a variety of molecular pathways.(32, 33) One potential mechanism involves accumulation of somatic variation, observed to increase with age, (34, 35) and thought to reflect a decline in mitochondrial replicative quality control.(36) The recent characterisation of nucleotide constraint across the mitochondrial genome (37) provides a mechanism for quantifying the overall burden of mitochondrial genome variation in an individual, or mitochondrial genome ‘quality’. This offers a conceptual tool to measure changes indicative of progressive age-related processes.

To evaluate both monogenic MD risk and broader age-related pathology, we employed the Medical Genome Reference Bank (MGRB), a biobank of healthy older Australians with bigenomic sequencing data and clinical data incorporating metabolic parameters, cognitive and physical function, as well as mood and quality of life measures.(35) We have established the frequency of pathogenic nDNA and mtDNA MD variants in this population, with as many as 1 in 60 individuals carrying a pathogenic mtDNA MD variant with (with heteroplasmy ≥1%), approximately 1 in 500 individuals carrying a pathogenic nDNA MD variant with autosomal dominant (AD) manifestations, and estimated autosomal recessive (AR) risk rates as high as 1 in 3,880 individuals. Furthermore, we have characterised mitochondrial constraint metrics, observing an increase in summed but not mean constraint with increasing age and total number of (non-haplotagging) mtDNA variants.

Therefore, this study aimed to address two critical points: 1) evaluating the association between carrying pathogenic mtDNA or AD nDNA MD variants and specific clinical features indicative of MD in healthy older individuals, and 2) investigating whether overall mitochondrial quality, quantified by mtDNA nucleotide constraint, is associated with indicators of aging and neurodegeneration.

## Methods

*Cohort*: the MGRB is a biobank of genetically characterised and deeply phenotyped healthy older Australians of predominantly European descent.(35) The cohort evaluated in this study included 2,794 participants drawn exclusively from the ASPREE study.(38–41) Individuals had lived to at least 75 years of age with no history of cancer, cardiovascular disease or dementia.(35)

*Genome sequencing*: the bigenomic whole genome sequencing (WGS) pipeline for this cohort has been detailed previously.(35) Nuclear variation data used in this study was extracted between January and April 2021 from the custom-built MGRB online Vectis platform (https://sgc.garvan.org.au/explore). Pathogenic or likely pathogenic variants with dominant inheritance arising in the MD gene set were included. Variants in mtDNA were called to a heteroplasmy cutoff of 1% using *mity* v0.4, an mtDNA variant analysis tool developed specifically for use with WGS data.(42) Pathogenic mtDNA variants were defined using Mitomap’s list of 130 confirmed pathogenic mtDNA variants (https://www.mitomap.org/MITOMAP, accessed 01/03/2025).(43)

*Mitochondrial haplogroups and constraint*: mitochondrial haplogroups and haplotagging single nucleotide polymorphisms (SNPs) were ascertained using Haplogrep v3.2.1.(44, 45) Mitochondrial nucleotide constraint scores were derived from mitochondrial local constraint values reported by Lake *et al*.(37) Summed constraint score (Summed CS) was calculated by summing constraint values for each mtDNA variant across an individual’s mitochondrial genome after removal of haplotagging SNPs. Mean constraint score (Mean CS) was calculated by dividing the Summed CS by the total number of variants contributing to that score.

*Clinical phenotypic measures*: clinical measures identified as MD-relevant phenotypic features that were available in the MGRB data set spanned four domains: cognitive function, physical (neuromuscular) function, endocrine-metabolic function, and mood. Clinical measures evaluated in each domain are summarised in **Supplementary Table 1** and included objective measures as well as health-related quality of life measures from the 12-item short form survey (SF-12). Hearing, vision and cardiac phenotype data, although strongly associated with MD, contained insufficient detail in the MGRB for evaluation in this study.

*MD variant associations*: as there were only one or a few (*n* = 1-5) of each specific variant identified in the cohort, analysis of mtDNA variant associations required grouping of variants by phenotypic manifestations available within the MGRB dataset: diabetes, renal, short stature, cognitive, stroke-like, myopathy, neuropathy. Clinical measures of interest for each phenotype were defined for analysis (**Supplementary Table 2**) and relevant clinical measures in phenotype variant carriers were compared to non-variant carriers. Pathogenic mtDNA variant carriers were also combined for analysis of relevant clinical measures compared to non-carriers. Analyses (a) using variant heteroplasmy threshold cutoffs of ≥2.5% and ≥5%, and (b) removing homoplasmic variants (>95%), were also undertaken. Variants in nDNA with monoallelic manifestations were too few to group by phenotype and were analysed in aggregate only.

*Statistical analyses*: age- and sex-adjusted regression models were used for association analyses; linear regression for continuous outcomes with heteroskedasticity robust standard errors and bias reduced (Firth penalised) logistic regression for binary outcomes. Model diagnostics indicated mild departures from normality for some bounded clinical scales (3MS and CES-D scores), consistent with ceiling and floor effects, respectively. Creatinine was log-transformed prior to analysis to improve residual normality and heteroskedasticity. Constraint scores (mean and summed CS, as standardised z-scores) were modelled as continuous predictors. Effect modification by sex was evaluated by including interaction terms between constraint scores and sex, and sex stratified analyses were performed. Welch *t*-tests and Fisher’s exact tests were undertaken for complementary unadjusted descriptive analyses. Adjustment for multiple comparison testing was made using a false discovery rate (FDR) correction, with significance defined at *q*<0.05. For phenotype-specific groups, additional exploratory analyses were undertaken to evaluate relationships between heteroplasmy and continuous outcome measures using Spearman rank correlation. Power was limited for phenotype-specific carrier groups with small sample counts, and detectable effect sizes were explored using simulation-based power analyses with R studio pwr package (see **Supplementary 02_Power**).

## Results

The average age across the cohort was 79.6 ± 3.4 years (range 75.3-93.8 years), with females representing 57% of subjects (1,599/2,794). Across the cohort, there were six nDNA variants identified which are associated with monoallelic disease manifestations, of which five are associated with multisystem manifestations (**Table 1**). A total of 56 individuals each carried one pathogenic mtDNA variant, equating to a prevalence of 1 in 50 healthy older individuals (56/2,794) carrying a pathogenic mtDNA variant. Within this group of 56 individuals, there were 32 distinct mtDNA variants represented (**Table 1**), with the majority of variants clustering at below 10% (42/56 (75%) <10% heteroplasmy) or above 95% (9/56, 16%) heteroplasmy, consistent with the bimodal heteroplasmy distribution observed for all mtDNA variation. (**Supplementary Figure 1**). Of the nine variants that were homoplasmic or near homoplasmic (heteroplasmy >95%), all were variants which clinically associate with isolated sensorineural hearing loss (SNHL) or Leber’s hereditary optic neuropathy (LHON). The nuclear and mitochondrial variants identified are summarised, together with relevant MGRB phenotypes for evaluation, in **Table 1**.

**Table 1a.** Nuclear DNA variants associated with monoallelic autosomal dominant manifestations identified in the MGRB cohort, with disease associations and relevant MGRB phenotype domains.

| Gene | Variant; consequence | Disease association(s) | Relevant MGRB phenotypes |
| --- | --- | --- | --- |
| *MFN2* | chr1-12069698-C-T;<br>NM_014874.4(*MFN2*):c.2119C>T (p.Arg707Trp) | CMT/HMSN-VI, MSL, optic neuropathy,<br>cognitive impairment | Cognitive, neuropathy |
| *OPA1* | chr3-193335054-T-A;<br>NM_130837.3(*OPA1*):c.536T>A (p.Leu179Ter) | Optic Atrophy; deafness, CPEO,<br>myopathy, ataxia | Myopathy |
| *OPA1* | chr3-193353231-C-T;<br>NM_130837.3(*OPA1*):c.868C>T (p.Arg290Ter) | Optic Atrophy; deafness, CPEO,<br>myopathy, ataxia | Myopathy |
| *TWNK* | chr10-102748875-G-A;<br>NM_021830.5(*TWNK*):c.908G>A (p.Arg303Gln) | CPEO; myopathy, ataxia, cardiomyopathy,<br>neuropathy, endocrinopathy, depression | Cognitive, endocrinopathy, myopathy, neuropathy,<br>seizures |
| *POLRMT* | chr19-620488-C-G;<br>NM_005035.4(*POLRMT*):c.2641-1G>C | CPEO; developmental delay, hypotonia,<br>short stature, intellectual disability | Cognitive, endocrinopathy, myopathy, seizures |
| *CYCS* | chr7-25163747-C-CTCTTTT;<br>NM_018947.6(*CYCS*):c.-8-2_-8-1insAAAAGA, p.? | Thrombocytopenia | N/A |

CMT Charcot-Marie-Tooth, CPEO chronic progressive external ophthalmoplegia, HMSN-VI hereditary motor and sensory neuropathy-6, MGRB Medical Genome Reference Bank, MSL multiple symmetric lipomatosis, N/A not available

**Table 1b.**
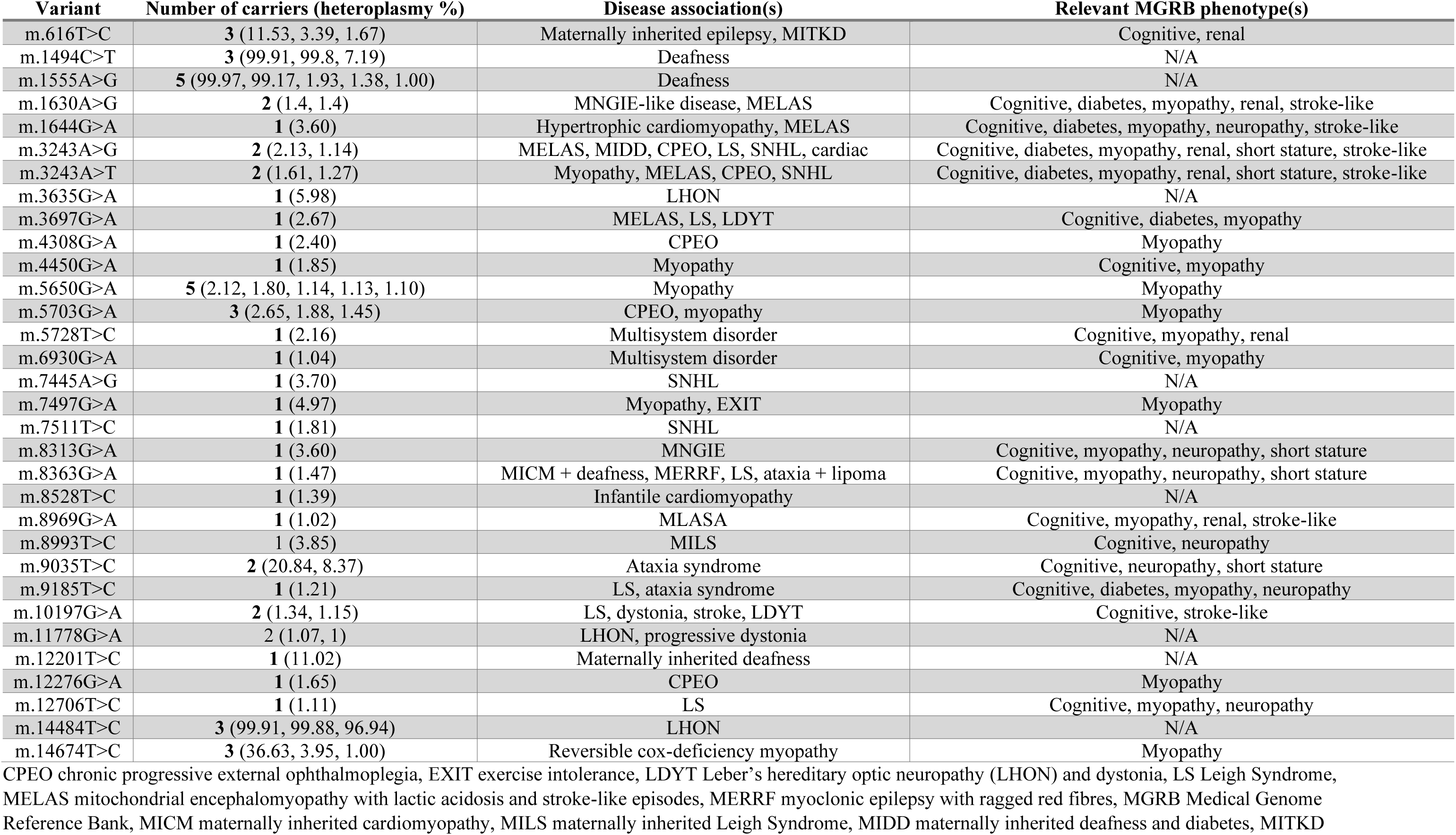
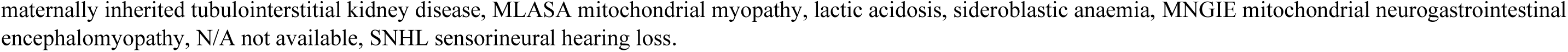
Mitochondrial DNA variants identified in the MGRB cohort, with disease associations and relevant MGRB phenotype domains

| Variant | Number of carriers (heteroplasmy %) | Disease association(s) | Relevant MGRB phenotype(s) |
| --- | --- | --- | --- |
| m.616T>C | 3 (11.53, 3.39, 1.67) | Maternally inherited epilepsy, MITKD | Cognitive, renal |
| m.1494C>T | 3 (99.91, 99.8, 7.19) | Deafness | N/A |
| m.1555A>G | 5 (99.97, 99.17, 1.93, 1.38, 1.00) | Deafness | N/A |
| m.1630A>G | 2 (1.4, 1.4) | MNGIE-like disease, MELAS | Cognitive, diabetes, myopathy, renal, stroke-like |
| m.1644G>A | 1 (3.60) | Hypertrophic cardiomyopathy, MELAS | Cognitive, diabetes, myopathy, neuropathy, stroke-like |
| m.3243A>G | 2 (2.13, 1.14) | MELAS, MIDD, CPEO, LS, SNHL, cardiac | Cognitive, diabetes, myopathy, renal, short stature, stroke-like |
| m.3243A>T | 2 (1.61, 1.27) | Myopathy, MELAS, CPEO, SNHL | Cognitive, diabetes, myopathy, renal, short stature, stroke-like |
| m.3635G>A | 1 (5.98) | LHON | N/A |
| m.3697G>A | 1 (2.67) | MELAS, LS, LDYT | Cognitive, diabetes, myopathy |
| m.4308G>A | 1 (2.40) | CPEO | Myopathy |
| m.4450G>A | 1 (1.85) | Myopathy | Cognitive, myopathy |
| m.5650G>A | 5 (2.12, 1.80, 1.14, 1.13, 1.10) | Myopathy | Myopathy |
| m.5703G>A | 3 (2.65, 1.88, 1.45) | CPEO, myopathy | Myopathy |
| m.5728T>C | 1 (2.16) | Multisystem disorder | Cognitive, myopathy, renal |
| m.6930G>A | 1 (1.04) | Multisystem disorder | Cognitive, myopathy |
| m.7445A>G | 1 (3.70) | SNHL | N/A |
| m.7497G>A | 1 (4.97) | Myopathy, EXIT | Myopathy |
| m.7511T>C | 1 (1.81) | SNHL | N/A |
| m.8313G>A | 1 (3.60) | MNGIE | Cognitive, myopathy, neuropathy, short stature |
| m.8363G>A | 1 (1.47) | MICM + deafness, MERRF, LS, ataxia + lipoma | Cognitive, myopathy, neuropathy, short stature |
| m.8528T>C | 1 (1.39) | Infantile cardiomyopathy | N/A |
| m.8969G>A | 1 (1.02) | MLASA | Cognitive, myopathy, renal, stroke-like |
| m.8993T>C | 1 (3.85) | MILS | Cognitive, neuropathy |
| m.9035T>C | 2 (20.84, 8.37) | Ataxia syndrome | Cognitive, neuropathy, short stature |
| m.9185T>C | 1 (1.21) | LS, ataxia syndrome | Cognitive, diabetes, myopathy, neuropathy |
| m.10197G>A | 2 (1.34, 1.15) | LS, dystonia, stroke, LDYT | Cognitive, stroke-like |
| m.11778G>A | 2 (1.07, 1) | LHON, progressive dystonia | N/A |
| m.12201T>C | 1 (11.02) | Maternally inherited deafness | N/A |
| m.12276G>A | 1 (1.65) | CPEO | Myopathy |
| m.12706T>C | 1 (1.11) | LS | Cognitive, myopathy, neuropathy |
| m.14484T>C | 3 (99.91, 99.88, 96.94) | LHON | N/A |
| m.14674T>C | 3 (36.63, 3.95, 1.00) | Reversible cox-deficiency myopathy | Myopathy |

CPEO chronic progressive external ophthalmoplegia, EXIT exercise intolerance, LDYT Leber's hereditary optic neuropathy (LHON) and dystonia, LS Leigh Syndrome, MELAS mitochondrial encephalomyopathy with lactic acidosis and stroke-like episodes, MERRF myoclonic epilepsy with ragged red fibres, MGRB Medical Genome Reference Bank, MICM maternally inherited cardiomyopathy, MILS maternally inherited Leigh Syndrome, MIDD maternally inherited deafness and diabetes, MITKD
maternally inherited tubulointerstitial kidney disease, MLASA mitochondrial myopathy, lactic acidosis, sideroblastic anaemia, MNGIE mitochondrial neurogastrointestinal encephalomyopathy, N/A not available, SNHL sensorineural hearing loss.

### Phenotype associations

When evaluating for phenotypes associating with mtDNA variants, no significant associations for the specified outcomes measures were identified for phenotype variant carriers compared to non-carriers (**Table 2**). Amongst carriers of variants associated with diabetes (*n* = 9; all heteroplasmy 1-4%), none had a diagnosis of diabetes (odds ratio OR 0.68, 95% CI 0.01-5.41), and there was no significant difference in blood glucose between groups (mean difference 0.14; 95% confidence interval −0.22-0.50). Whilst a positive correlation between variant heteroplasmy and blood glucose levels was observed, as observed by Cannon *et al.*,(26) carrier numbers were small and heteroplasmy levels low, and findings were not statistically significant (data not shown). For carriers of variants associated with renal dysfunction (*n* = 11; heteroplasmy range 1-12%), there was no significant difference in odds of chronic kidney disease (CKD; OR 0.29; 95% CI 0.03-1.24) or hypertension (OR 0.73; 95% CI 0.22-2.96), and mean creatinine was not significantly different after adjusting for age and sex (% difference −2.36; 95% CI −8.50-4.21) when compared to non-carriers. Height did not differ significantly in carriers (*n* = 8; heteroplasmy range 1-21%) of variants associated with short stature (mean difference 0.02; 95% CI −0.02-0.05) and BMI showed no significant difference between groups (mean difference −0.84; 95% CI −4.40-2.71). No strokes occurred in carriers of variants associated with stroke-like events (*n* = 10; heteroplasmy range 1-4%) and there was no difference in odds of stroke between groups (OR 1.61; 95% CI 0.01-12.87).

**Table 2.**
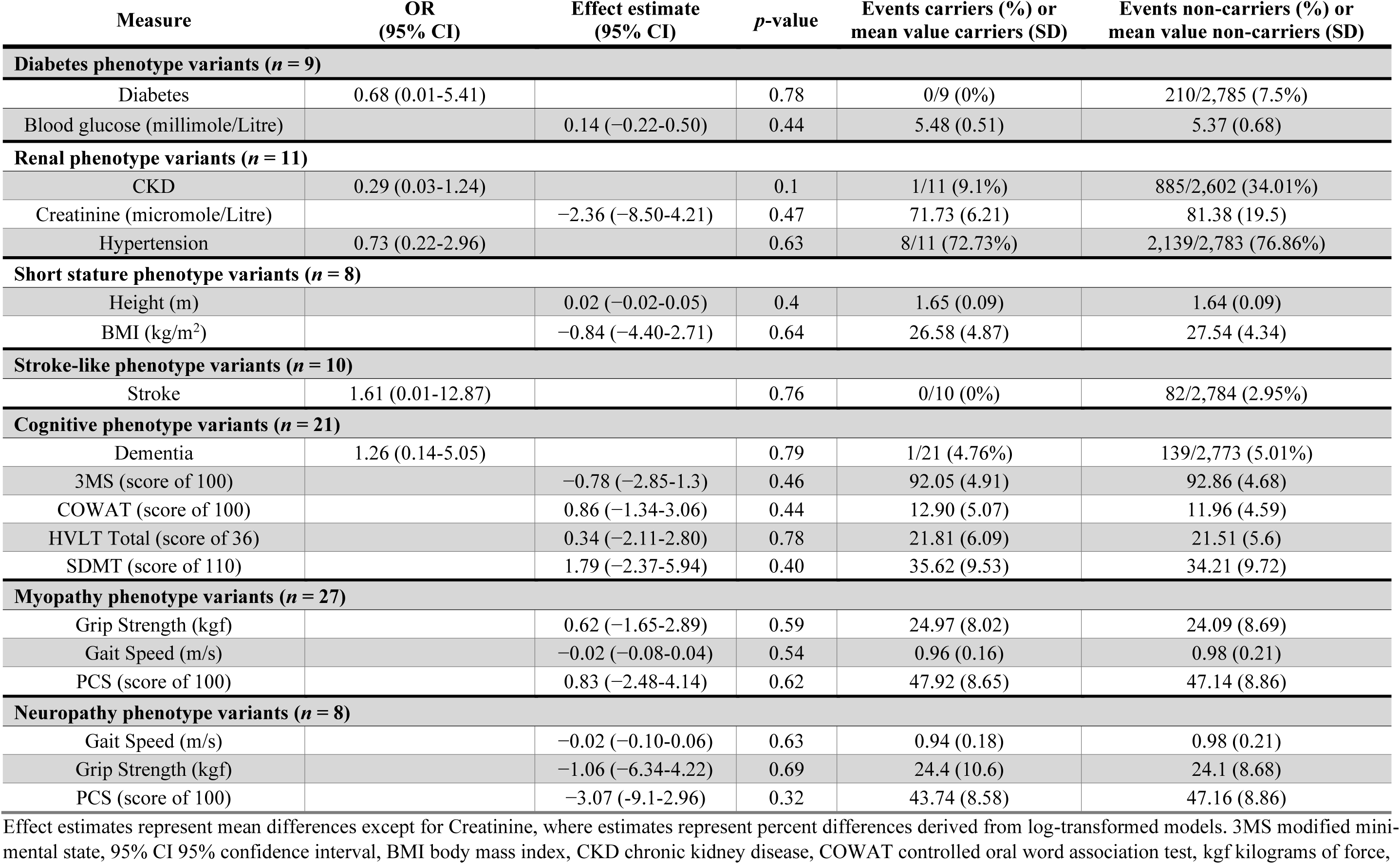
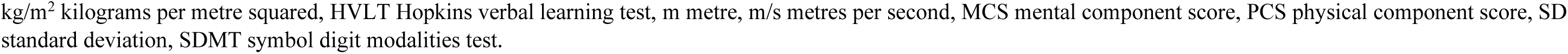
Comparison of relevant phenotypic measures between variant carriers and non-carriers.

| Measure | OR<br>(95% CI) | Effect estimate<br>(95% CI) | p-value | Events carriers (%) or<br>mean value carriers (SD) | Events non-carriers (%) or<br>mean value non-carriers (SD) |
| --- | --- | --- | --- | --- | --- |
| **Diabetes phenotype variants (n = 9)** |  |  |  |  |  |
| Diabetes | 0.68 (0.01-5.41) |  | 0.78 | 0/9 (0%) | 210/2,785 (7.5%) |
| Blood glucose (millimole/Litre) |  | 0.14 (−0.22-0.50) | 0.44 | 5.48 (0.51) | 5.37 (0.68) |
| **Renal phenotype variants (n = 11)** |  |  |  |  |  |
| CKD | 0.29 (0.03-1.24) |  | 0.1 | 1/11 (9.1%) | 885/2,602 (34.01%) |
| Creatinine (micromole/Litre) |  | −2.36 (−8.50-4.21) | 0.47 | 71.73 (6.21) | 81.38 (19.5) |
| Hypertension | 0.73 (0.22-2.96) |  | 0.63 | 8/11 (72.73%) | 2,139/2,783 (76.86%) |
| **Short stature phenotype variants (n = 8)** |  |  |  |  |  |
| Height (m) |  | 0.02 (−0.02-0.05) | 0.4 | 1.65 (0.09) | 1.64 (0.09) |
| BMI (kg/m<sup>2</sup>) |  | −0.84 (−4.40-2.71) | 0.64 | 26.58 (4.87) | 27.54 (4.34) |
| **Stroke-like phenotype variants (n = 10)** |  |  |  |  |  |
| Stroke | 1.61 (0.01-12.87) |  | 0.76 | 0/10 (0%) | 82/2,784 (2.95%) |
| **Cognitive phenotype variants (n = 21)** |  |  |  |  |  |
| Dementia | 1.26 (0.14-5.05) |  | 0.79 | 1/21 (4.76%) | 139/2,773 (5.01%) |
| 3MS (score of 100) |  | −0.78 (−2.85-1.3) | 0.46 | 92.05 (4.91) | 92.86 (4.68) |
| COWAT (score of 100) |  | 0.86 (−1.34-3.06) | 0.44 | 12.90 (5.07) | 11.96 (4.59) |
| HVLT Total (score of 36) |  | 0.34 (−2.11-2.80) | 0.78 | 21.81 (6.09) | 21.51 (5.6) |
| SDMT (score of 110) |  | 1.79 (−2.37-5.94) | 0.40 | 35.62 (9.53) | 34.21 (9.72) |
| **Myopathy phenotype variants (n = 27)** |  |  |  |  |  |
| Grip Strength (kgf) |  | 0.62 (−1.65-2.89) | 0.59 | 24.97 (8.02) | 24.09 (8.69) |
| Gait Speed (m/s) |  | −0.02 (−0.08-0.04) | 0.54 | 0.96 (0.16) | 0.98 (0.21) |
| PCS (score of 100) |  | 0.83 (−2.48-4.14) | 0.62 | 47.92 (8.65) | 47.14 (8.86) |
| **Neuropathy phenotype variants (n = 8)** |  |  |  |  |  |
| Gait Speed (m/s) |  | −0.02 (−0.10-0.06) | 0.63 | 0.94 (0.18) | 0.98 (0.21) |
| Grip Strength (kgf) |  | −1.06 (−6.34-4.22) | 0.69 | 24.4 (10.6) | 24.1 (8.68) |
| PCS (score of 100) |  | −3.07 (−9.1-2.96) | 0.32 | 43.74 (8.58) | 47.16 (8.86) |

Effect estimates represent mean differences except for Creatinine, where estimates represent percent differences derived from log-transformed models. 3MS modified mini-mental state, 95% CI 95% confidence interval, BMI body mass index, CKD chronic kidney disease, COWAT controlled oral word association test, kgf kilograms of force,
kg/m<sup>2</sup> kilograms per metre squared, HVLT Hopkins verbal learning test, m metre, m/s metres per second, MCS mental component score, PCS physical component score, SD standard deviation, SDMT symbol digit modalities test.

A larger number of identified variants had potential to affect cognitive function (*n* = 21; heteroplasmy range 1-21%) providing greater power to detect a small effect. However, proportions of dementia amongst variant carriers were similar to non-carriers (4.76% vs 5.01%; OR 1.26; 95% CI 0.14-5.05), and there were no significant differences in specific cognitive assessment scores including modified mini mental state (3MS; mean difference −0.78; 95% CI −2.85-1.3), controlled oral word association test (COWAT; mean difference 0.86; 95% CI −1.34-3.06), Hopkins verbal learning test (HVLT) total score (mean difference 0.34, 95% CI −2.11-2.80) or symbol-digit modalities test (SDMT; mean difference 1.79; 95% CI −2.37-5.94). Overall, no significant differences were detected in physical function metrics between carriers of neuropathy-associated variants (*n* = 8; heteroplasmy range 1-21%) and myopathy-associated variants (*n* = 27; heteroplasmy range 1-37%), although the direction of association was consistently negative for physical outcomes in neuropathy variant carriers. Specifically, mean differences for gait speed (myopathy −0.02; 95% CI −0.08-0.04; neuropathy −0.02; 95% CI −0.10-0.06), grip strength (myopathy 0.62; 95% CI −1.65-2.89; neuropathy −1.06; 95% CI −6.34-4.22) and physical component scores (PCS) (myopathy 0.83; 95% CI −2.48-4.14; neuropathy −3.07; 95% CI −9.1-2.96) were all non-significant.

### Associations of combined pathogenic mtDNA variants

Combining all pathogenic mtDNA variant carriers (*n* = 56) for analysis of outcome measures compared to non-carriers (**Table 3**; **Figure 1**), surprisingly suggested an improved mood domain in variant carriers. This was represented by significantly lower CES-D scores (mean difference −0.9; 95% CI −1.49-−0.31; *p* 0.003) and better self-reported mental health reflected by higher mental component scores (MCS; mean difference 1.83; 95% CI −0.27-3.40; *p* = 0.02) in variant carriers, and a trend towards reduced odds of depression (OR 0.57; 95% CI 0.30-1.03; *p* = 0.06). Although directionally concordant across the domain, effect sizes were small and, after adjusting for multiple testing, were non-significant and remained so for odds of depression.

**Figure 1.**
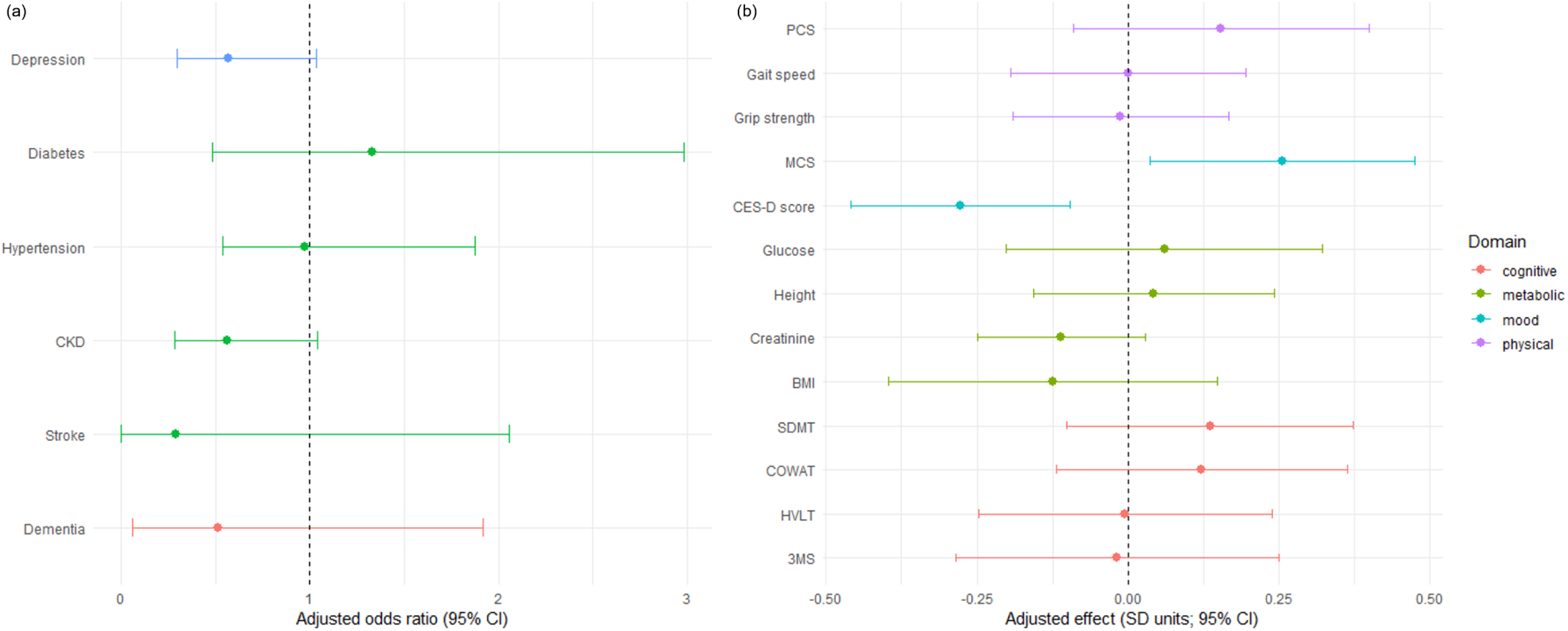
Phenotypic associations of mtDNA variant carriers. Adjusted odds ratio (OR) for binary measures and adjusted effect (SD units) for continuous measures for MGRB cohort mtDNA variant carriers vs non-carriers **(a)** associations between carriage of any mtDNA variant and binary measures (adjusted OR), grouped by phenotypic domain and **(b)** associations between carriage of any mtDNA variant and standardised continuous measures (adjusted effect), grouped by phenotypic domain. 3MS Modified Mini Mental Status, BMI body mass index, CES-D centre for epidemiological studies-depression, CKD chronic kidney disease, COWAT controlled oral word association test, HVLT Hopkins Verbal Learning Test, MCS mental component score, PCS physical component score, SDMT symbol digit modalities score

**Table 3.**
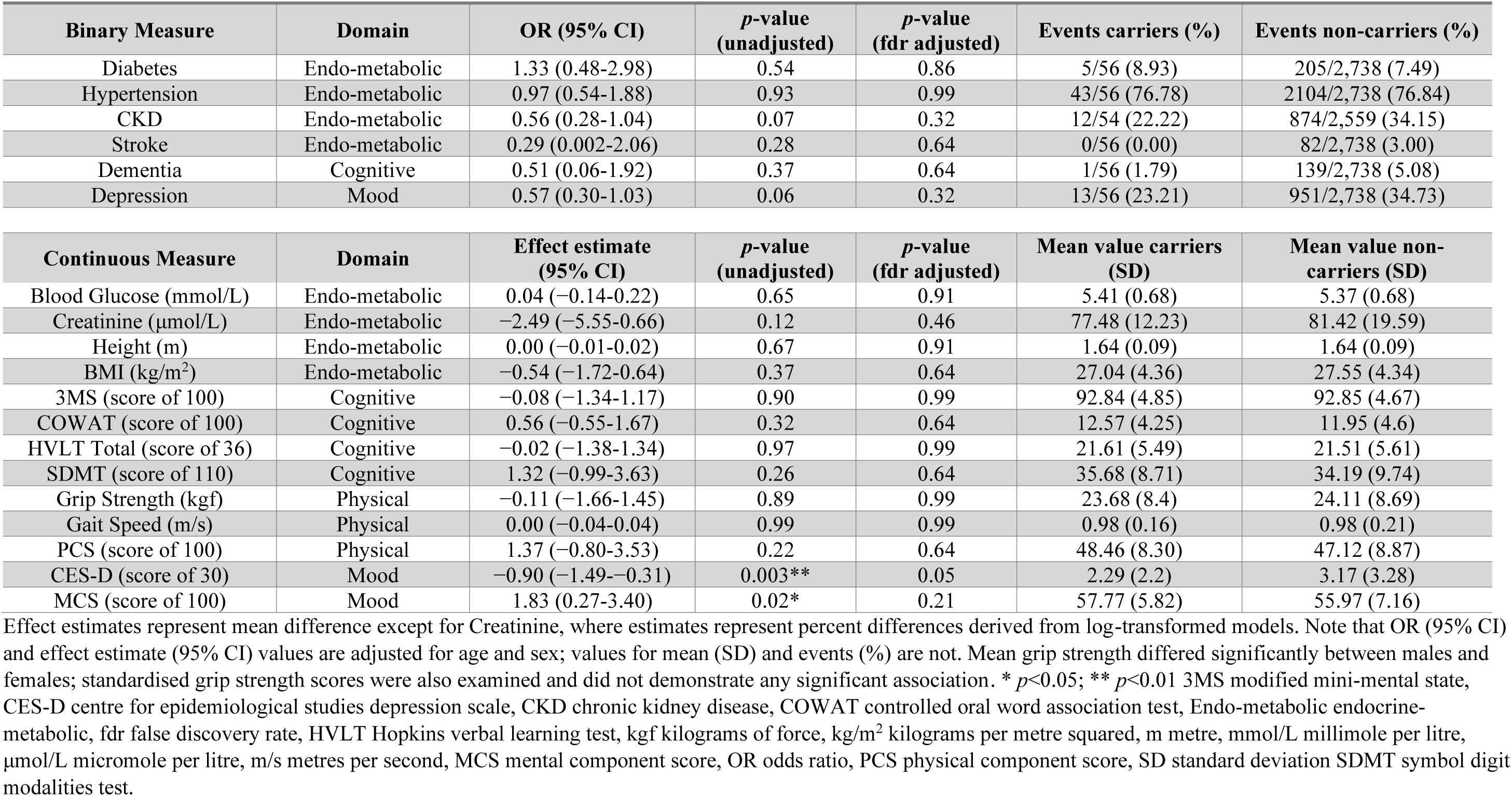
Analysis of phenotype associations for all mtDNA variant carriers, compared to non-carriers.

Mean difference in gait speed (0.00; 95% CI −0.04-0.04), grip strength (−0.11; 95% CI −1.66-1.45), blood glucose (0.04; 95% CI −0.14-0.22), height (0.00; 95% CI −0.01-0.02), 3MS (−0.08; 95% CI −1.34- 1.17) and HVLT total scores (−0.02; 95% CI −1.38-1.34) were closely comparable between carriers and non-carriers (**Figure 1**). BMI (mean difference −0.54; 95% CI −1.72-0.64) and creatinine (% difference −2.49; 95% CI −5.55-0.66) were relatively lower in variant carriers but this difference was not significant. However, when considered alongside odds of CKD (OR 0.56; 95% CI 0.28-1.04), these measurements, although not significantly reduced, may be indicative of comparatively lower muscle mass.(46)

Increasing the heteroplasmy threshold for pathogenic mtDNA variants to 2.5% (*n* = 26) or 5% (*n* = 18) did not reveal any significant associations (**Supplementary Table 3**), although increased the magnitude of effect on CES-D score. Interestingly, removing homoplasmic variants (>95%; all associated with single-organ manifestations of isolated hearing or vision loss) from the analysis increased the observed effects for MCS, CES-D scores, depression and CKD. However, after adjusting for multiple testing, they were non-significant or remained so for depression (**Supplementary Table 3**).

### nDNA variant associations

Evaluating nDNA variants (*n* = 6, all heterozygous) indicated significantly better 3MS scores in nDNA variant carriers compared to non-carriers (mean difference 2.85, 95% CI 1.5-4.21; adj *p* = 0.00003, adj *p* = 0.0005). However, COWAT scores associated negatively with variant carriers (mean difference −3.47, 95% CI −6.67-−0.28, *p* = 0.03, adj *p* = 0.22) and there were no significant differences in other cognitive domain measures including odds of dementia, HVLT and SDMT scores. There were no other significant associations after adjusting for multiple testing (**Supplementary Table 3**). Incorporation of AD nDNA MD variant carriers with combined mtDNA variant carriers had no impact on associations (**Supplementary Table 3**).

### Associations with mitochondrial constraint measures

Summed and mean mitochondrial local CS were calculated for each sample after removal of haplotagging variants. Individual samples contained a median of 18 variants (IQR 10-23). Summed CS ranged from 0.01 to 27.33 (mean 4.36 and SD 3.14). Summed CS were observed to correlate closely to the number of variants (Pearson’s r = 0.899, *p* <0.001) and increased slightly with age (Pearson’s r = 0.22, *p* < 0.001). Mean CS ranged from 0.01 to 0.70 (mean 0.24 and SD 0.09). Mean CS did not increase with number of variants (Pearson’s r = -0.003, *p* = 0.865) and increased minimally with age (Pearson’s r = 0.048, *p* = 0.011) (**Supplementary Figures 1 and 2**). The mean number of variants and summed CS did not differ significantly between pathogenic mtDNA variant carriers and non-carriers, however, mean CS was significantly higher in mtDNA variant carriers (0.27 compared to 0.24; mean difference 0.03; 95% CI 0.01-0.06, *p* = 0.01).

**Figure 2.**
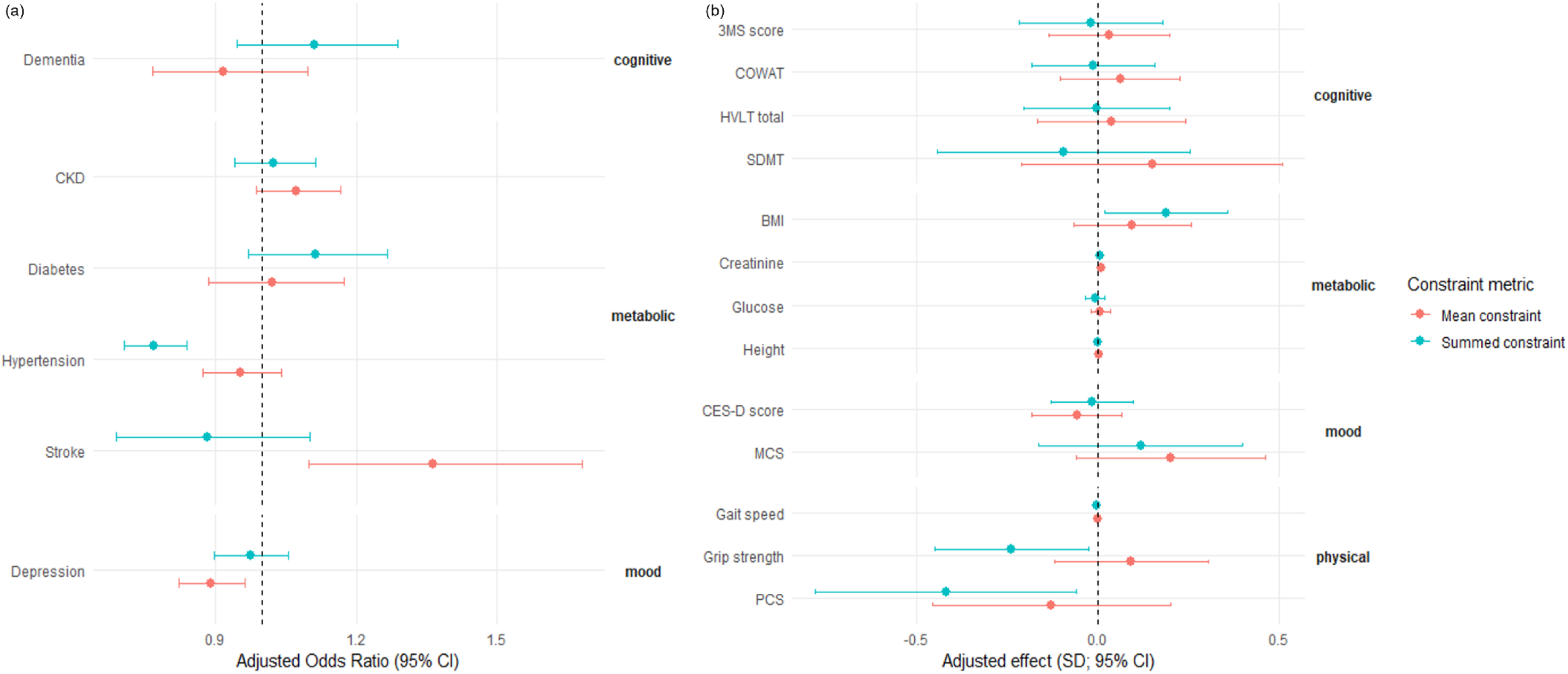
Associations of summed and mean mitochondrial constraint scores (salmon and teal respectively) with domain-specific binary and continuous measures of aging **(a)** associations between standardised mean and summed constraint scores and binary measures (adjusted odds ratio), grouped by phenotypic domain and **(b)** associations between standardised mean and summed constraint scores and continuous measures (adjusted effect), grouped by phenotypic domain. 3MS Modified Mini Mental Status, BMI body mass index, CES-D centre for epidemiological studies-depression, CKD chronic kidney disease, COWAT controlled oral word association test, HVLT Hopkins Verbal Learning Test, MCS mental component score, PCS physical component score, SDMT symbol digit modalities score.

When comparing mitochondrial constraint with phenotypic measures (**Table 4** and **Figure 2**), there were no significant associations with individual cognitive scores or odds of dementia, or with glucose, diabetes, BMI or CKD. The odds of hypertension reduced significantly with increasing Summed CS (OR 0.77 per unit of SD; 95% CI 0.71-0.84), including after adjustment for multiple testing (*p* = 8.8*10^-8^). With increasing Mean CS, creatinine increased (% difference 0.85 per SD unit; 95% CI 0.11-1.60; *p* = 0.02), however this small change was unlikely of clinical significance and did not remain significant after adjustment for multiple testing. Odds of stroke increased (OR 1.36 per SD unit; 95% CI 1.10 to 1.68, *p* = 0.004) and odds of depression decreased (OR 0.89 per SD unit; 95% CI 0.82-0.96; *p* = 0.004). However, these findings were not consistent within their respective domains across binary and continuous outcomes and did not remain significant after adjustment for multiple testing (**Table 4**).

**Table 4.** Associations of standardised mean and summed mitochondrial constraint scores with aging-related measures in the MGRB.

| Binary Measure | Domain | Summed Constraint<br>OR (95% CI) | p-value<br>(unadjusted) | p-value<br>(fdr adjusted) | Mean Constraint<br>OR (95% CI) | p-value<br>(unadjusted) | p-value<br>(fdr adjusted) |
| --- | --- | --- | --- | --- | --- | --- | --- |
| Diabetes | Endo-metabolic | 1.11 (0.97-1.27) | 0.11 | 0.43 | 1.02 (0.89-1.17) | 0.78 | 0.88 |
| Hypertension | Endo-metabolic | 0.77 (0.71-0.84) | 2.3*10<sup>-9</sup>*** | 8.8*10<sup>-8</sup>*** | 0.85 (0.87-1.04) | 0.28 | 0.67 |
| CKD | Endo-metabolic | 1.02 (0.94-1.11) | 0.58 | 0.81 | 1.07 (0.99-1.17) | 0.10 | 0.43 |
| Stroke | Endo-metabolic | 0.88 (0.69-1.10) | 0.30 | 0.67 | 1.36 (1.10-1.68) | 0.004** | 0.05 |
| Dementia | Cognitive | 1.11 (0.95-1.30) | 0.18 | 0.56 | 0.92 (0.77-1.10) | 0.35 | 0.72 |
| Depression | Mood | 0.97 (0.90-1.06) | 0.54 | 0.81 | 0.89 (0.82-0.96) | 0.004** | 0.05 |

| Continuous Measure | Domain | Summed Constraint<br>Effect estimate (95% CI) | p-value<br>(unadjusted) | p-value<br>(fdr adjusted) | Mean Constraint<br>Mean difference (95% CI) | p-value<br>(unadjusted) | p-value<br>(fdr adjusted) |
| --- | --- | --- | --- | --- | --- | --- | --- |
| Blood Glucose<br>(millimole/Litre) | Endo-metabolic | -0.01 (-0.03-0.02) | 0.59 | 0.81 | 0.01 (-0.02-0.03) | 0.62 | 0.81 |
| Creatinine (micromole/Litre) | Endo-metabolic | 0.61 (-0.13-1.36) | 0.10 | 0.34 | 0.85 (0.11-1.60) | 0.02* | 0.11 |
| Height (m) | Endo-metabolic | -0.0002 (-0.002-0.002) | 0.87 | 0.90 | 0.001 (-0.001-0.004) | 0.19 | 0.56 |
| BMI (kg/m<sup>2</sup>) | Endo-metabolic | 0.19 (-0.02-0.36) | 0.03* | 0.16 | 0.09 (-0.07-0.26) | 0.25 | 0.67 |
| 3MS (score of 100) | Cognitive | -0.02 (-0.22-0.18) | 0.84 | 0.90 | 0.03 (-0.13-0.20) | 0.72 | 0.86 |
| COWAT (score of 100) | Cognitive | -0.01 (-0.18-0.16) | 0.87 | 0.90 | 0.06 (-0.10-0.22) | 0.56 | 0.74 |
| HVLT Total (score of 36) | Cognitive | 0.003 (-0.20-0.20) | 0.97 | 0.97 | 0.04 (-0.17-0.24) | 0.71 | 0.86 |
| SDMT (score of 110) | Cognitive | -0.09 (-0.44-0.25) | 0.59 | 0.81 | 0.15 (-0.21-0.51) | 0.42 | 0.72 |
| Grip Strength (kgf) | Physical | -0.24 (-0.45--0.03) | 0.03* | 0.16 | 0.09 (-0.12-0.30) | 0.40 | 0.72 |
| Gait Speed (m/s) | Physical | -0.004 (-0.01-0.003) | 0.27 | 0.67 | -0.002 (-0.01-0.01) | 0.66 | 0.84 |
| PCS (score of 100) | Physical | -0.42 (-0.78--0.06) | 0.02* | 0.16 | -0.13 (-0.46-0.20) | 0.45 | 0.74 |
| CES-D (score of 30) | Mood | -0.02 (-0.13-0.10) | 0.78 | 0.88 | -0.06 (-0.18-0.07) | 0.36 | 0.72 |
| MCS (score of 100) | Mood | 0.12 (-0.16-0.40) | 0.41 | 0.72 | 0.20 (-0.06-0.46) | 0.13 | 0.45 |

Effect estimates represent mean difference except for Creatinine, where estimates represent percent differences derived from log-transformed models. \* $p < 0.05$ ; \*\* $p < 0.01$ ; \*\*\* $p < 0.001$ . 3MS modified mini-mental state, CES-D centre for epidemiological studies depression scale, CKD chronic kidney disease, COWAT controlled oral word association test, Endo-metabolic endocrine and/or metabolic, fdr false discovery rate, HVLT Hopkins verbal learning test, kgf kilograms of force, kg/m<sup>2</sup> kilograms per metre squared, m metre, m/s metres per second, MCS mental component score, MGRB medical genome reference bank, OR odds ratio, PCS physical component score, SD standard deviation SDMT symbol digit modalities test.

There was an apparent inverse association between physical measures and Summed CS. This was reflected in significantly lower mean grip strength (mean difference −0.24 kilograms of force per SD unit; 95% CI −0.45-−0.03; *p* = 0.03) and self-reported physical health represented by lower PCS (mean difference −0.42 per SD unit; 95% CI −0.78-−0.06; *p* = 0.02) with increasing Summed CS. However, gait speed was not significantly reduced (mean difference −0.004 m/s per SD unit; 95% CI −0.01-0.003, *p* = 0.27) and these associations did not remain significant after adjusting for multiple testing. Inclusion of a sex interaction term and sex stratified analyses did not alter these findings.

## Discussion

Pathogenic MD variants are common in the population,(18–22, 25, 26) yet there is limited data evaluating the clinical relevance of such variants for carriers in unselected populations. This study, conducted in an “extreme phenotype” healthy older individual biobank,(35) demonstrates that individuals carrying a range of pathogenic mtDNA MD variants and AD nDNA MD variants, can live in good health through to old age with no indication of adverse impacts across a range of clinically relevant and functionally important measures. Our evaluation incorporates the use of multiple detailed cognitive measures, as well as sensitive objective measures of physical function in older age, and subjective quality of life measures. These important domains have not previously been evaluated in carriers of pathogenic MD variants, and in conjunction with data from clinical cohorts, add valuable context for clinical counselling and management.

Amongst clinical outcomes of relevance and available to study in the MGRB population, there was no clear indication of mild variant-related phenotypes below the threshold for clinical detection, arguing against a mild or subclinical spectrum of disease in this cohort. In particular, evaluation of physical and cognitive function by a range of measures demonstrated no association with pathogenic mitochondrial variation. However, individual phenotype groups were small, limiting minimum detectable effects and therefore a small to moderate effect cannot be excluded. Examining larger cohorts across a range of ages and relevant phenotypes (such as individuals with diabetes or renal disease) will be necessary to further the understanding of variant penetrance and expression. In particular, careful examination of relevant mild phenotypes such as vision and hearing loss, outcomes not assessable in this cohort, will be important. Whilst not statistically significant, several observations warrant consideration: amongst neuropathy variant carriers, the direction of association across all physical domain measures was negative. Although none of these individual associations were significant, the small number of carriers in this group (*n* = 8) limited power to detect a small to moderate effect, and further evaluation in larger cohorts – especially amongst older variant carriers – would be of value. Contrary to expectations, and observations of individuals with MD,(47) combined evaluation of all mtDNA variants indicated that mental health and mood may be enhanced in MD variant carriers, particularly heteroplasmic variants that typically associate with multiorgan manifestations, by both subjective and objective measure. Conceivably, such an effect may render mtDNA variants more persistent in populations than predicted based on their deleterious mitochondrial effects. However, further study in larger populations is required to explore this observation. Finally, with the observation of relatively lower BMI, creatinine, and CKD in variant carriers, further evaluation of biomarkers, particularly those associated with muscle manifestations (such as FGF21), may be of interest.(48–50)

Taken together, the findings in this study support variable penetrance of MD variants between unselected individuals and clinical cohorts, as was also observed by Cannon and colleagues examining UK Biobank data.(26) However, several factors need to be considered in interpreting these results. For mtDNA variants, the tissue sampled and heteroplasmy threshold effect are relevant,(2, 51) with studies often demonstrating increasing probability and severity of manifestations with increasing heteroplasmy load. Yet, manifest disease with low heteroplasmy also occurs.(52–54) Variant heteroplasmy varies by tissue, and genetic data in this study were derived exclusively from blood DNA, which may have lower variant heteroplasmy than other (especially disease affected) tissues.(54) Approximately half of the variants identified here had heteroplasmy <2.5%. Whilst low heteroplasmy variants are less likely to cause disease (below threshold), a single blood heteroplasmy measure does not capture what may be manifest in disease-relevant tissues.(54, 55) Nevertheless, the presence of a pathogenic variant, regardless of blood heteroplasmy, confers potential risks to an individual and to subsequent generations in a maternal lineage. Analysis with heteroplasmy thresholds set at 2.5% or 5% did not show any change in associations. Interestingly, exclusion of homoplasmic variants (associated with isolated single-organ vision or hearing manifestations) appeared to increase the observed effects on mood and MCS, although not to statistical significance after correction for multiple testing. It is also important to consider that somatic variants (which accumulate from the age of 60-70) and nuclear mitochondrial transcripts (NUMTs) may contaminate pathogenic variants as both may confound low heteroplasmy variant calling. Comparison of mtDNA variation in the MGRB with other cohorts including gnomAD and UK BioBank (not shown), indicates variant findings are comparable to those reported in these larger cohorts, and the additional (presumed somatic) variant burden with age was small compared to observed mtDNA variation overall. The analysis paradigm also presumes single variant effects, whilst multiple factors may be relevant to variant penetrance and clinical manifestations, including elements such as haplogroup, nDNA-mtDNA interactions and epigenetic modifications,(56) which were not encompassed in this analysis.

For AD nDNA variants identified in the MGRB population, applying penetrance estimates from the literature would imply far higher rates of population AD nDNA MD risk (around 1 in 600) than encountered in routine clinical practice. The data presented here, although limited in number, does not demonstrate negative associations for variant carriers. This supports the hypothesis that variants identified in clinically unselected carriers may have differential implications compared to those identified through established diagnostic or family cascade testing, a pattern corroborated by studies in other contexts.(29–31, 57) This underscores the complementary nature of evaluation from both clinical cohorts and population-based studies, acknowledging the uncertainty between these two ends of a conceptual spectrum, where potential mild and/or single organ disease may not be recognised as associated with MD.

The MGRB is a comparatively small cohort, limiting power to detect smaller changes. A similar study in the UK Biobank (26) found significant association of diabetes with m.3243A>G variants, especially with increasing heteroplasmy >10%. There were only 2 carriers of the m.3243A>G variant in the current study, and heteroplasmy for both was <2.5%. In this context, the absence of an association is not unexpected. Conversely, manifest disease has been observed in association with blood heteroplasmy below 1%,(58) underscoring the challenge of examining rare variation across a heteroplasmy spectrum from a non-disease relevant tissue in a smaller cohort. Clinical manifestations of particular relevance to MD, including hearing and vision, could not be examined in this cohort due to the insufficiently overlapping subset of individuals with hearing or vision data. Seizures and cardiac outcomes relevant to MD were also not able to be confidently ascertained from MGRB data and were therefore not evaluated. These would be of most relevance to assess in future studies of larger representative cohorts.

Mitochondrial genome constraint has recently been modelled (37) and has the potential to provide a conceptual avenue to quantify “quality” across an individual’s mitochondrial genome and for comparison between individuals. In this study, there was suggestion that summed mitochondrial genome constraint may be associated with reduced physical (muscle) function whilst mean constraint was not. Although not significant after adjusting for multiple testing, this is an interesting observation as summed constraint may more accurately model cumulative mtDNA variant burden in post-mitotic tissues such as muscle, compared to high-turnover tissues like blood which are subject to stronger selection pressures. Summed CS associated inversely with odds of hypertension which was unexpected given the evolving evidence linking mitochondrial dysfunction with hypertension and its sequelae.(59) This inverse association may help explain the apparent divergence in odds of stroke when comparing summed versus mean CS respectively. Further elucidation of this association may shed light on the mechanisms linking mitochondrial dysfunction, hypertension and vascular risk. Surprisingly, there was no indication that cognitive dysfunction associated with constraint, suggesting that mitochondrial constraint measures may not capture the association between mitochondrial dysfunction and neurodegeneration. Nevertheless, mitochondrial constraint metrics are an inviting theoretical possibility with some interesting observations in a small, older cohort suggesting potential utility, especially for measures of frailty. Consideration of constraint metrics in additional relevant tissues, across varied ages and disease states including MD, in larger cohorts, and with refinements such as heteroplasmy weighting may yield informative results.

## Conclusion

Pathogenic variants associated with MD are common in the population and may be carried through to old age in good health without indication of mild or subclinical mitochondrial disease. MD variants identified incidentally in unselected individuals may not have the same associations, penetrance, or risk as those identified in the clinical context. Measures of mitochondrial genome constraint may offer potential as a biomarker for aging and frailty and warrant further evaluation and refinement in larger cohorts and, where possible, varied tissues and contexts.

## Supporting information

Supplementary Power

## Declarations

### Ethics

The ASPREE Biobank study was approved by the Alfred Hospital Human Research Ethics Committee, and the study was performed in accordance with the ethical standards in the Declaration of Helsinki.

### Data Availability

The MGRB genetic data used for this study is available on request at https://sgc.garvan.org.au/terms/mgrb/index.html.

Access to clinical data for ASPREE participants may be considered on application https://aspree.org/aus/.

## Supplementary Materials

Supplementary 1_figures

Supplementary 2_power

## Author Contribution

E.W. conceptualisation, data curation, formal analysis, funding acquisition, investigation, methodology, validation, visualisation, writing – original draft and review & editing

G.Q. data curation, writing – review and editing

S.R. data curation, resources and software, writing – review & editing

M.H. data curation, software, writing – review & editing

J.C. data curation, resources, software, writing – review & editing

C.Y. data curation, software, writing – review and editing

S.K. resources, software, writing – review & editing

C.L. conceptualisation, supervision, validation, visualisation, writing – review & editing

P.L. conceptualisation, investigation, resources, writing – review & editing

R.L.D. conceptualisation, methodology, administration, supervision, validation, visualisation, writing – review & editing

C.M.S. conceptualisation, funding acquisition, methodology, administration, resources, supervision, validation, writing – review & editing

## Use of AI statement

During preparation of this work, the authors used Google Gemma-3n-e4b AI Model on LM Studio to assist with editing. After using this tool, the authors reviewed and edited the content as needed, and take full responsibility for the content of the publication.

