## Supplementary Tables for "Mitochondrial Disease variation in healthy older adults: a genotype-phenotype assessment linking pathogenic variants and mitochondrial constraint"

**Supplementary Table 1**: Phenotypic domains and clinical measures evaluated by domain.

| Domain | Measure(s) | Variable Type | Result; range |
| --- | --- | --- | --- |
| Cognitive | Dementia  3MS Score  HVLT Total  COWAT  SDMT | Binary  Continuous  Continuous  Continuous  Continuous | Present / Absent  Points (out of 100)  Points (out of 36)  Points (out of 100)  Points (out of 110) |
| Physical / Neuromuscular | Gait Speed (3metre gait)  Grip Strength  PCS (from SF-12) | Continuous  Continuous  Continuous | Speed (metres/second)  Strength (kilograms of force)  Standardised score (out of 100) |
| Endocrine-Metabolic | Diabetes  Hypertension  CKD  Stroke  Height  BMI  Creatinine  Blood Glucose | Binary  Binary  Binary  Binary  Continuous  Continuous  Continuous  Continuous | Present / Absent  Present / Absent  Present / Absent  Present / Absent  metres  metres/kilogram^2^  micromoles/litre  millimoles/litre |
| Mood | Depression  CES-D score  MCS (from SF-12) | Binary  Continuous  Continuous | Present / Absent  Points (out of 100)  Standardised score (out of 100) |

3MS modified mini mental state, BMI Body Mass Index, CES-D Centre for epidemiologic studies depression scale, CKD chronic kidney disease, COWAT controlled oral word association test, HVLT Hopkins verbal learning test, MCS mental component score, PCS physical component score, SDMT symbol digit modalities test, SF-12 12-item short form health-related quality of life survey.

**Supplementary Table 2**: Phenotype groups and outcome measures evaluated.

| Phenotype | carriers (*n*) | Domain | Primary MEASURE(s) | Secondary MEASURE(s) |
| --- | --- | --- | --- | --- |
| Diabetes | 9 | Endocrine-metabolic | Diabetes | Blood Glucose |
| Renal | 11 | Endocrine-metabolic | CKD | Serum Creatinine  Hypertension |
| Short Stature | 8 | Endocrine-metabolic | Height | BMI |
| Cognitive | 21 | Cognitive | Dementia  3MS | COWAT  HVLT Total  SDMT |
| Stroke-like | 10 | Cognitive | Stroke |  |
| Myopathy | 27 | Physical | Grip Strength | Gait Speed  PCS |
| Neuropathy | 8 | Physical | Gait Speed | Grip Strength  PCS |

3MS modified mini mental state, BMI body mass index, CKD chronic kidney disease, COWAT controlled oral word association test, HVLT Hopkins verbal learning test, PCS physical component score, SDMT symbol digit modalities test.

**Supplementary table 3** Analysis of AD nDNA variants, mitochondrial disease variants with adjusted heteroplasmy thresholds, and any MD variants

| Binary measures | | | |  |  | | |  | |
| --- | --- | --- | --- | --- | --- | --- | --- | --- | --- |
| Binary Measure | AD nDNA variants  OR (95% CI) | | mtDNA heteroplasmy >5%  OR (95% CI) | | | | mtDNA heteroplasmic variants only  OR (95% CI) | | Any MD variants  (nDNA or mtDNA) |
| Diabetes | 0.84 (0.01-7.16) | | 1.95 (0.39-6.34) | | | | 1.60 (0.58-3.62) | | 1.18 (0.43-2.62) |
| Hypertension | 0.33 (0.07-1.54) | | 1.42 (0.49-5.45) | | | | 0.85 (0.45-1.69) | | 0.86 (0.49-1.56) |
| CKD | 0.18 (0.00-1.68) | | 1.21 (0.44-3.06) | | | | 0.48 (0.22-0.96) *  (*p* = 0.04; adj *p* = 0.23) | | 0.51 (0.26-0.92) *  (*p* = 0.03; adj *p* = 0.16) |
| Stroke | 2.73 (0.02-24.86) | | 1.12 (0.01-8.39) | | | | 0.33 (0.00-2.37) | | 0.26 (0.00-1.86) |
| Dementia | 1.52 (0.01-13.58) | | 0.62 (0.00-4.63) | | | | 0.59 (0.07-2.23) | | 0.46 (0.05-1.73) |
| Depression | 1.28 (0.22-5.81) | | 0.75 (0.25-1.94) | | | | 0.52 (0.25-1.00) | | 0.61 (0.33-1.07) |
| Continuous Measure | AD nDNA variants  Effect estimate (95% CI) | | mtDNA heteroplasmy >5%  Effect estimate (95% CI) | | | | mtDNA heteroplasmic variants only  Effect estimate (95% CI) | | Any MD variants  (nDNA or mtDNA) |
| Blood Glucose (millimole/Litre) | −0.22 (−0.83-0.4) | | 0.09 (−0.22-0.39) | | | | 0.02 (−0.17-0.21) | | 0.02 (−0.15-0.19) |
| Creatinine (micromole/Litre) | 1.72 (−7.92-12.38) | | −1.36 (−6.24-3.77) | | | | −3.04 (−6.37-0.40) | | −2.16 (−5.07-0.84) |
| Height (metres) | 0.03 (−0.01-0.08) | | 0.01 (−0.02-0.04) | | | | 0.01 (−0.01-0.03) | | 0.01 (−0.01-0.02) |
| BMI (kilogram per metre^2^) | −1.25 (−2.77-0.28) | | −0.64 (−3.01-1.74) | | | | − 0.71 (−2.00-0.59) | | −0.61 (−1.69-0.46) |
| 3MS (score of 100) | 2.85 (1.5-4.21)***  (*p* = 0.00003; adj *p* = 0.0007) | | 1.64 (−0.20-3.48) | | | | − 0.43(−1.81-0.95) | | 0.21 (−0.95-1.36) |
| COWAT (score of 100) | −3.47 (−6.67—0.28)*  (*p* = 0.03; adj *p* = 0.31) | | 0.09 (−1.56-1.73) | | | | 0.84 (−0.39-2.07) | | 0.16 (−0.91-1.24) |
| HVLT Total (score of 36) | 0.39 (−3.71-4.49) | | 0.33 (−1.99-2.65) | | | | −0.53 (−2.03-0.96) | | 0.02 (−1.26-1.29) |
| SDMT (score of 110) | 5.99 (−1.22-13.20) | | −0.68 (−4.88-3.52) | | | | 1.90 (−0.74-4.55) | | 1.79 (−0.41-3.99) |
| Grip Strength (kilograms of force) | 4.85 (−2.18-11.88) | | −0.15 (−3.2-2.91) | | | | 0.14 (−1.51-1.79) | | 0.39 (−1.18-1.96) |
| Gait Speed (metres per second) | −0.08 (−0.17-−0.00)*  (*p* = 0.05; adj *p* = 0.31) | | 0.01 (−0.07-0.08) | | | | −0.01 (−0.05-0.04) | | −0.01 (−0.05-0.03) |
| PCS (score of 100) | −2.25 (−9.08-4.57) | | 1.52 (−2.44-5.47) | | | | 1.29 (−0.99-3.58) | | 1.02 (−1.04-3.07) |
| CES-D (score of 30) | 0.49 (−3.51-−4.49) | | −1.23 (−2.05-−0.41)**  (*p* = 0.003; adj *p* = 0.06) | | | | −0.94 (−1.60-−0.27) **  (*p =* 0.006; adj *p* = 0.06) | | −0.77 (−1.41-−0.13)*  (*p* = 0.02; adj *p* = 0.16) |
| MCS (score of 100) | 0.98 (−5.73-7.69) | | 0.14 (−2.8-3.07) | | | | 2.35 (0.66-4.05) **  (*p* = 0.007; adj *p* = 0.06) | | 1.76 (0.23-3.28)*  (*p* = 0.02; adj *p* = 0.16) |

Effect estimates represent mean difference except for Creatinine, where estimates represent percent differences derived from log-transformed models. mtDNA heteroplasmy >5% = mtDNA variants implementing a minimum 5% heteroplasmy threshold; mtDNA heteroplasmic variants only = mtDNA variants with heteroplasmy >1% but excluding homoplasmic variants (>95%; with isolated single-organ manifestations of vision or hearing loss); Any MD variants (nDNA or mtDNA) = including both mtDNA variants (heteroplasmy >1%) and AD nDNA variants. * *p*<0.05; ***p*<0.01; *** *p* <0.001. 3MS modified mini-mental state, adj adjusted, CES-D centre for epidemiological studies depression scale, CI confidence interval, CKD chronic kidney disease, COWAT controlled oral word association test, HVLT Hopkins verbal learning test, MCS mental component score, PCS physical component score, SDMT symbol digit modalities test.
