## Supplementary Figures for "Mitochondrial Disease variation in healthy older adults: a genotype-phenotype assessment linking pathogenic variants and mitochondrial constraint"

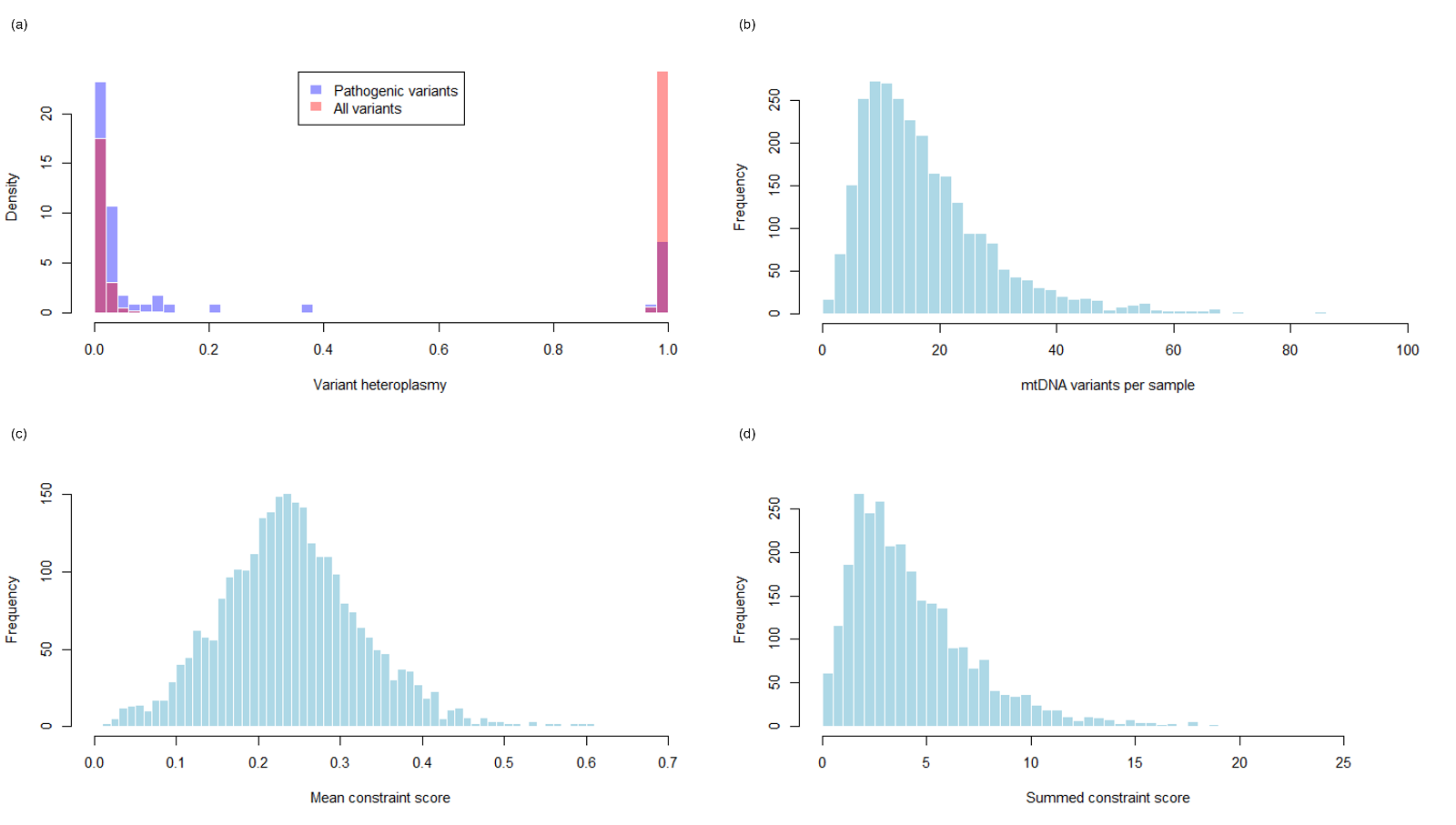

**Supplementary Figure 1** demonstrating distribution of (a) heteroplamy of all variation and pathogenic variation in the cohort, (b) mtDNA variation by sample, (c) mean and (d) summed constraint scores

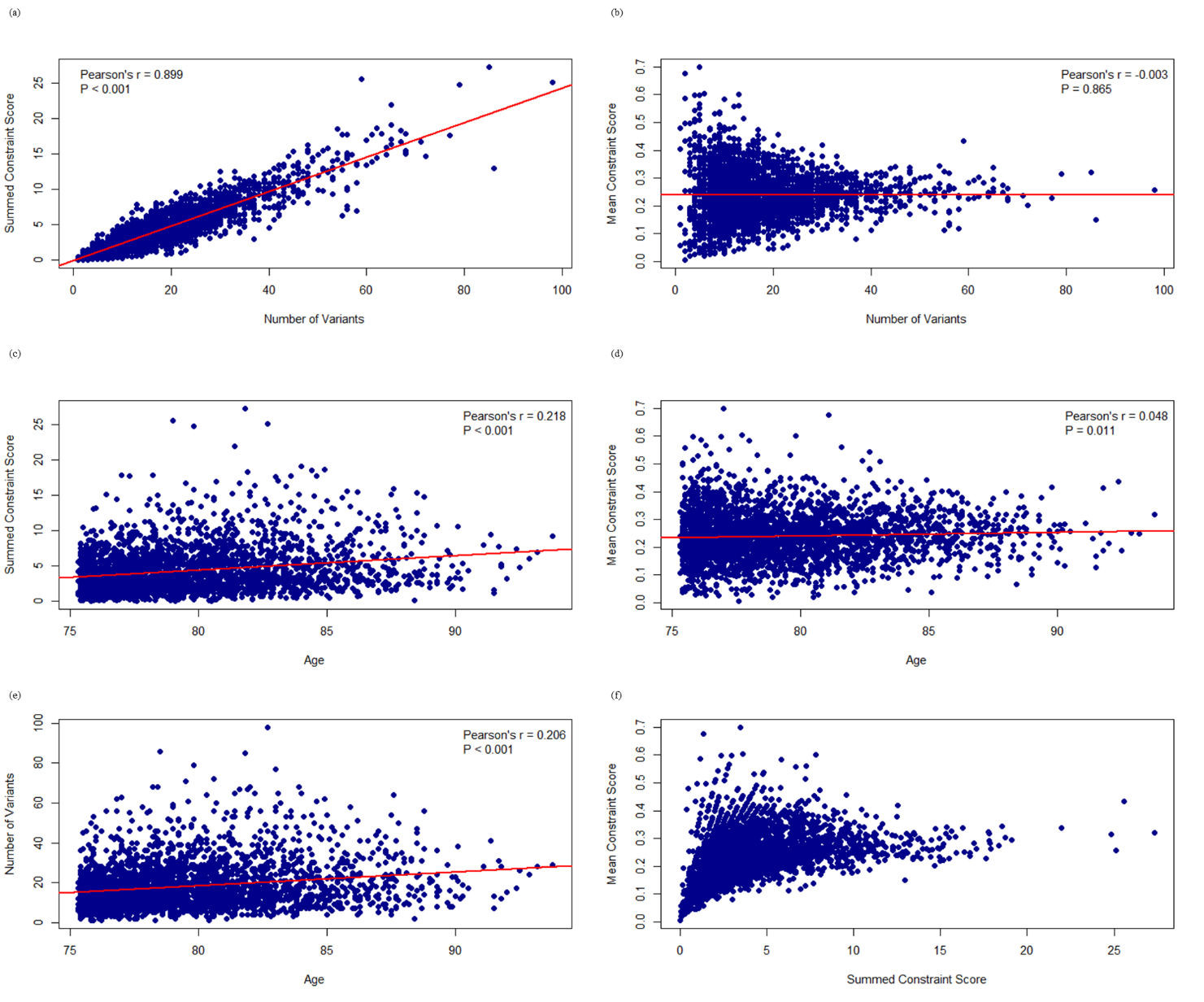

**Supplementary figure 2** demonstrating (a) close relationship between summed constraint score (Summed CS) and number of variants per sample, whilst (b) mean constraint score (Mean CS) does not change with number of variants. (c) shows positive correlation between Summed CS and age, whereas (d) Mean CS minimally increases with age. The relationship between number of variants and age is shown in (e) and that between Mean CS and Summed CS is shown in (f).
